# Evaluating the Potential of Large Language Models for Vestibular Rehabilitation Education: A Comparison of ChatGPT, Google Gemini, and Clinicians

**DOI:** 10.1101/2024.01.24.24301737

**Authors:** Yael Arbel, Yoav Gimmon, Liora Shmueli

## Abstract

**Objective:** We aimed to evaluate the performance of two publicly available large language models, ChatGPT and Google Gemini in response to multiple-choice questions related to vestibular rehabilitation.

**Methods:** The study was conducted among 30 physical therapist professionals experienced with VR (vestibular rehabilitation) and 30 physical therapy students. They were asked to complete a Vestibular Knowledge Test (VKT) consisting of 20 multiple-choice questions that were divided into three categories: (1) Clinical Knowledge, (2) Basic Clinical Practice, and (3) Clinical Reasoning. ChatGPT and Google Gemini were tasked with answering the same 20 VKT questions. Three board-certified otoneurologists independently evaluated the accuracy of each response using a 4-level scale, ranging from comprehensive to completely incorrect.

**Results:** ChatGPT outperformed Google Gemini with a 70% score on the VKT test, while Gemini scored 60%. Both excelled in Clinical Knowledge with a perfect score of 100% but struggled in Clinical Reasoning with ChatGPT scoring 50% and Gemini scoring 25%. According to three otoneurologic experts, ChatGPT’s accuracy was considered comprehensive in 45% of the 20 questions, while 25% were found to be completely incorrect. ChatGPT provided comprehensive responses in 50% of Clinical Knowledge and Basic Clinical Practice questions, but only 25% in Clinical Reasoning.

**Conclusion:** Caution is advised when using ChatGPT and Google Gemini due to their limited accuracy in clinical reasoning. While they provide accurate responses concerning Clinical Knowledge, their reliance on web information may lead to inconsistencies. ChatGPT performed better than Gemini. Healthcare professionals should carefully formulate questions and be aware of the potential influence of the online prevalence of information on ChatGPT’s and Google Gemini’s responses. Combining clinical expertise and clinical guidelines with ChatGPT and Google Gemini can maximize benefits while mitigating limitations.

**Impact Statement:** This study highlights the potential utility of large language models like ChatGPT in supplementing clinical knowledge for physical therapists, while underscoring the need for caution in domains requiring complex clinical reasoning. The findings emphasize the importance of integrating technological tools carefully with human expertise to enhance patient care and rehabilitation outcomes.

## Introduction

The integration of artificial intelligence (AI) technologies in healthcare has created a new era in patient care, marked by the exploration of the potential integration of AI chatbots, including ChatGPT (Generative Pre-trained Transformer) and Google Gemini, in medicine as a means to assist healthcare professionals. However, AI’s use in vestibular rehabilitation (VR), which requires precise knowledge and expertise, has not yet been thoroughly investigated. Understanding the capabilities of the language models (LLMs) and their potential applications in VR is vital for advancing evidence-based practices and delivering accurate knowledge to patients. This study, therefore, sought to assess the accuracy and grading of the information provided by ChatGPT concerning vestibular physiology and vestibular rehabilitation.

VR is a specialized form of treatment for patients suffering from dizziness. This exercise-based treatment program is designed to promote vestibular adaptation and substitution. Since its advent in the 1940s,^1^ VR has undergone significant advancements and is now the recommended therapy for dizziness, vestibular dysfunction, and benign paroxysmal positional vertigo (BPPV).^2^ The Barany Society, has amended its definitions for diagnosing vestibular disorders several times since 2009.^3–6^ In addition, updated guidelines provide evidence-based recommendations on how to improve diagnostic accuracy and treatment efficiency.^2,7^ Implementing these guidelines is challenging for many clinicians and requires continuous learning to ensure consistent, evidence-based treatment and standards in VR. ^8,9^ In summary, VR is an evolving field requiring clinicians to continuously update their knowledge and research to provide evidence-based, effective treatment.

As VR advances, we aimed to explore how AI technologies like ChatGPT and Google Gemini can enhance access to knowledge for VR therapists. Effective use of AI can assist VR therapists in accessing knowledge. ChatGPT, an interactive chatbot powered by the GPT3.5 architecture developed by OpenAI, is a LLM that has demonstrated tremendous potential in accelerating learning and knowledge acquisition in diverse medical fields.^10^ Trained on a vast dataset from the Internet, ChatGPT excels at generating human-like responses in conversations and prompts across multiple languages and subject domains, making it a valuable tool for various applications.^11–13^

Recent studies have evaluated the accuracy level of the answers provided by various version of ChatGPT to knowledge-based questions in the field of medicine, both with respect to multiple-choice questions and to open-ended questions. When answering multiple-choice questions related to well-defined, established medical questionnaires, ChatGPT’s accuracy rate was found to vary across different domains, ranging from 42% in the field of ophthalmology, as measured by the OKAP (Ophthalmic Knowledge Assessment Program) test, to 76% in cardiology, specifically in the American Heart Association (AHA) and Advanced Cardiovascular Life Support (ACLS) exams.^11,12^

A more recent entry, Google’s Gemini, demonstrates significant potential as a conversational AI model, with extensive applications across industries such as customer service, healthcare, and education.^14^ Research compared the accuracy and precision of ChatGPT-3.5, ChatGPT-4, and Google Gemini in analyzing retinal detachment cases before surgery. The results showed that ChatGPT performed better than Google Gemini.^15^

The studies that assessed the accuracy and reproducibility of ChatGPT responses to open-ended questions involved grading the responses by several clinical domain experts using a scale that included four categories: (1) comprehensive, (2) correct but inadequate, (3) some correct and some incorrect, and (4) completely incorrect. For instance, a study on Bariatric Surgery^16^ found that the model provided "comprehensive" responses to 131 out of 151 questions, resulting in an accuracy rate of 86.8%. In a similar study regarding cirrhosis and hepatocellular carcinoma, an accuracy rate of 76.9% was determined.^17^

An alternative approach implemented by Gilson et al^18^ to determine the degree of accuracy on the AMBOSS Student Medical exam entailed evaluating the text output of each ChatGPT response across 3 qualitative metrics: logical justification of the answer selected, presence of information internal to the question, and presence of information external to the question.

The field of VR is constantly evolving, requiring individuals to engage in continuous learning to stay up-to-date. This study aims to investigate the use of AI in disseminating knowledge and to evaluate clinicians’ utilization of AI tools in clinical settings. We compared the performance of two publicly available LLMs, ChatGPT and Google Gemini, with that of physical therapists who received training in VR and that of physical therapy students. The comparison was based on answering multiple-choice questions. Based on the responses to the questionnaire, we concentrated on the responses of the best-performing language models and evaluated the accuracy of the explanations of these chosen answers by the language modal.

## Materials and Methods

### Data source, VKT multiple choice Questionnaire

In May 2023, a Vestibular Knowledge Test (VKT) consisting of 20 multiple-choice questions (one correct answer and three incorrect options (distractors) was developed (See supplemental Appendix 1). It was categorized into three groups: Clinical Knowledge, Basic Clinical Practice, and Clinical Reasoning. The test was validated through an anonymous online survey using Qualtrics.^19^ The survey link was distributed via WhatsApp, targeting two distinct groups of participants: 1) Physical therapists who had previously received training in VR, and 2) physical therapy students across all years of the degree. During the survey, before each question, appeared the prompt "Please select the correct answer.”

Out of the 60 respondents (30 Physical therapists and 30 physical therapy students) who completed the survey, 53% (n = 32) identified as female. The ages of the participants ranged from 21 to 51 years (M = 31.65, SD = 7.92). With respect to the 30 Physical therapists, the median duration of work experience as a Physical therapist was 11.08 years (range <2–23 years), and the median time since their last vestibular training was 2.44 years (range <0–12 years).

To assess the internal consistency of the questionnaire, reliability statistics were calculated using Cronbach’s alpha. For the physical therapy students, the Cronbach’s alpha value was α = .37, indicating relatively low internal consistency. In contrast, for the physical therapy group, the Cronbach’s alpha value was α = .68, suggesting a higher level of internal consistency.

### ChatGPT and Google Gemini Response Generation

To generate responses for VKT questions, we used the ChatGPT AI language model (2023 version) based on the GPT-3.5 architecture. This model was trained on data updated in September 2021. We also used Google Gemini (2024 version) to prompt the VKT questions. Both ChatGPT-3.5 and Google Gemini are freely available versions. We prompted the questions separately using the same chat in each language model.

### Grading

Two grading methods were implemented: (1) Multiple-choice questionnaire grading: A score of 1 was assigned to each multiple-choice question answered correctly, and a grade of 0 to an incorrect answer, resulting in a maximum total score of 20 points. We chose this technique to simulate human test-taking.

The second grading method was conducted to assess the accuracy of ChatGPT’s explanations, as it outperformed Google Gemini in the multiple-choice questions. (2) Question-response grading: A score was assigned to each explanation provided by ChatGPT, based on an independent assessment of its accuracy and compatibility with the answer provided. The review and grading of each response were independently performed by three board-certified otoneurologists who are experts in the vestibular field. The reviews were conducted based on evidence-based knowledge to determine the accuracy of the ChatGPT responses. The accuracy of each ChatGPT response was rated using the following scale: 1. “Comprehensive”: The answer is correct, and the explanation provided is accurate and comprehensive. 2. “Correct but inadequate”: Both the answer and the explanation are correct but not satisfactory in terms of completeness. 3. “Mixed with correct and incorrect/outdated data”: The answer is incorrect, and the explanation is partially accurate. 4. “Completely incorrect”: Both the answer and the explanation are incorrect. The accuracy of each answer was determined based on the median of the experts’ answers.

### Statistical analyses

The grading was determined by comparing ChatGPT’s and Google Gemini’s answers to the answer key. The model’s grading was determined by a single assessment. The means and standard deviation obtained on the 20 multiple-choice single-answer questions by the Physical therapists experienced in VR and physical therapy students with no prior knowledge of vestibular rehabilitation, were compared to the response points of the ChatGPT model and Google Gemini model. This was done using One Sample T-Test (test of means) to determine if there is a significant difference between response points earned between these respective groups. The results were then calculated as a total grade for each group and reported as percentages.

Response points from each category were compared between ChatGPT and Google Gemini by experienced VR physical therapists and physical therapy students. This comparison used a One Sample T-Test for each category (Clinical Knowledge, Basic Clinical Practice, and Clinical Reasoning), aiming to identify significant differences in response points across the groups for each category.The accuracy of the ChatGPT answers was rated by three experts and the score was determined according to the median of the three given scores. A correlation test was performed between both experts using Spearman analysis.

## Results

### Overall Performance

A VKT consisting of 20 multiple-choice single-answer questions was completed by 30 Physical therapists experienced in VR, 30 physical therapy students, ChatGPT-3.5 and Google Gemini (See supplemental Appendix 1). ChatGPT answered 70% of the questions correctly (14 out of 20), Google Gemini answered 60% of the questions correctly (12 out of 20), while the Physical therapists achieved a score of 76.3%, and the physical therapy students a score of 40.5% (Figure 1). A statistical analysis using t-tests revealed significant differences between the groups. When comparing overall performance, Physical therapists outscored ChatGPT and Google Gemini (t = 2.46, *p* < .05; t = 6.25, *p* < .001, respectively), with a large effect size (Cohen’s d = 2.89; Cohen’s d = 1.14, respectively). On the other hand, the performance of physical therapy students was significantly worse compared to ChatGPT and Google Gemini (t = -13.24, *p* < .001; t = -8.75, *p* < .001, respectively), with a large effect size (Cohen’s d = 2.44, Cohen’s d = 1.598, respectively).

**Figure 1:**
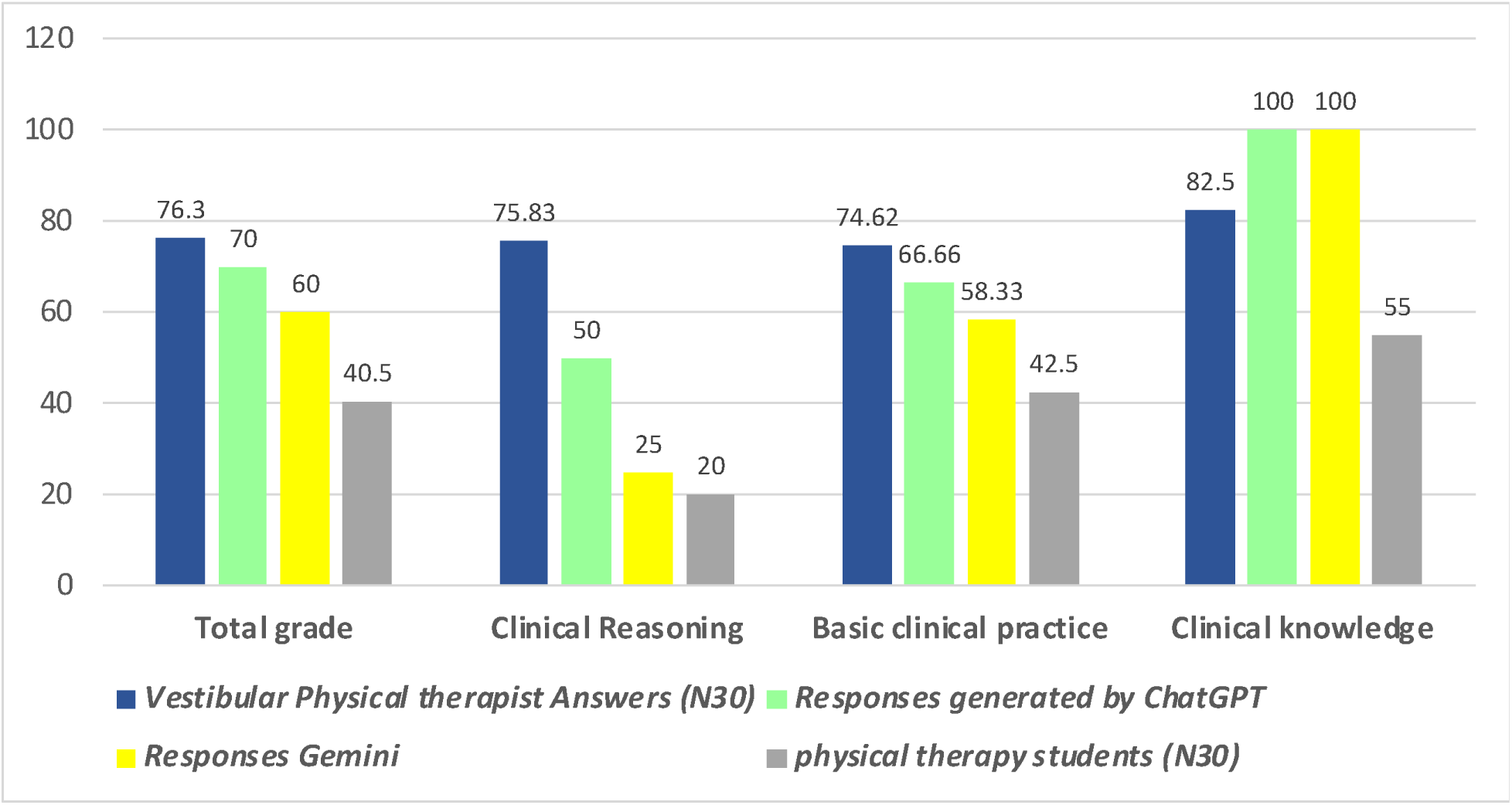
Performance of ChatGPT, Google Gemini, the Physical therapists and the physical therapy Students on the VKT, stratified by question type and topic.

### "Performance by Category Type"

A t-test analysis was conducted to compare performance in four categories of Clinical Knowledge, Basic Clinical Practice, and Clinical Reasoning among ChatGPT, Gemini, Physical therapists, and students. In Clinical Knowledge, both ChatGPT and Google Gemini answered all 4 questions correctly (100%). However, in the Clinical Reasoning category, ChatGPT managed to answer only 2 out of 4 correctly (50%), while Gemini answered only 1 correctly (25%). In the Basic Clinical Practice category, ChatGPT provided the correct answer to only 8 of the 12 questions (66.66%), outperforming Gemini, which answered 7 of the 12 questions correctly (58.33%).

In terms of clinical knowledge, Physical therapists and physical therapy students performed considerably worse than both ChatGPT and Gemini (t = 5.11, *p* < .001; t = - 10.25, *p* < .001, respectively), with a large effect size (Cohen’s d = 0.93; Cohen’s d = 1.87, respectively).

In Basic Clinical Practice category, Physical therapists significantly outperformed both ChatGPT (t = 2.83, *p* = .008 Cohen’s d =.51) with a medium effect size, and Google Gemini (t = 5.77, *p* < .001; Cohen’s d = 1.05) with large effect size. Conversely, physical therapy students performed significantly worse than both ChatGPT (t = -8.30, *p* < .001; Cohen’s d = 1.52) and Google Gemini (t = -5.47, *p* < .001; Cohen’s d = 1), with large effect sizes.

In the Clinical Reasoning category, Physical therapists significantly outperformed both ChatGPT (t = 5.86, *p* < .001; Cohen’s d = 1.07) and Google Gemini (t = 11.54, *p* < .001; Cohen’s d = 2.10) with large effect size. Conversely, physical therapy students performed significantly worse than ChatGPT (t = -7.10, p < .001; Cohen’s d = 1.29) with a large effect size, there was no significant difference in performance between physical therapy students and Google Gemini (t = -1.18, p = .246),

### Accuracy of the ChatGPT-generated explanations

The accuracy of ChatGPT’s response was assessed and categorized as either comprehensive, correct but inadequate, mixed (correct and incorrect/outdated data), and completely wrong. Based on the experts’ evaluation of the ChatGPT answers, 9 of the 20 answers were "comprehensive" (45%), 5 were “correct but inadequate” (25%), 1 was deemed “mixed with correct and incorrect responses” (5%) and the remaining 5 were considered "completely incorrect" (25%). When analyzed according to knowledge category, it was found that ChatGPT provides "comprehensive" responses to 2 of the 4 (50%) question related to Clinical Knowledge, to 6 of the 12 (50%) questions concerning "Basic Clinical Practice, and to only 1 of the 4 (25%) questions in the Clinical Reasoning category (Figure 2). Figure 3 illustrates further examples of ChatGPT prompts by category. ChatGPT performed in the test better than Gemini (that almost failed the knowledge test), therefore only the accuracy of the ChatGPT was evaluated by the experts.

**Figure 2:**
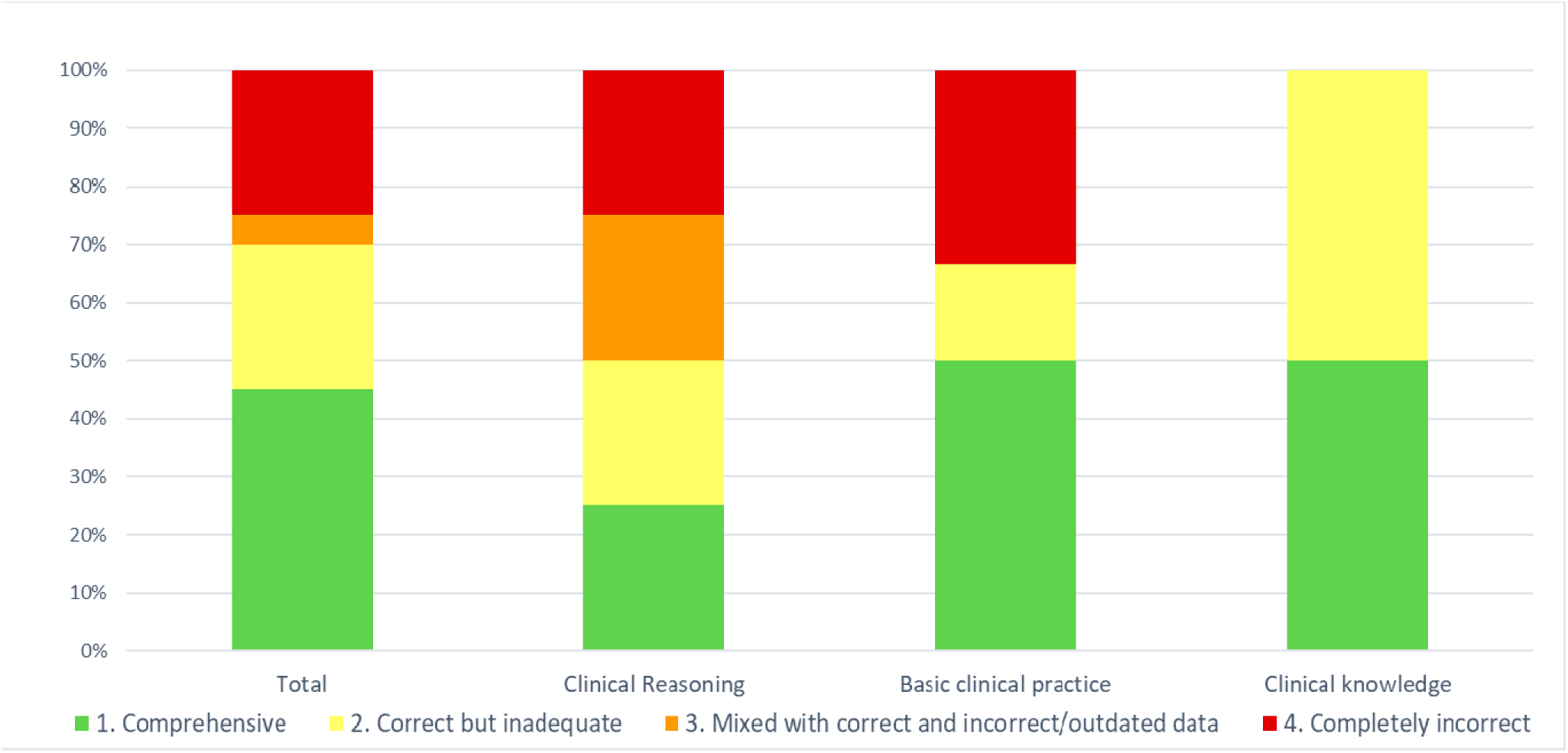
Accuracy of the responses generated by ChatGPT-3.5 to questions related to VR, categorized by knowledge category

**Figure 3:**
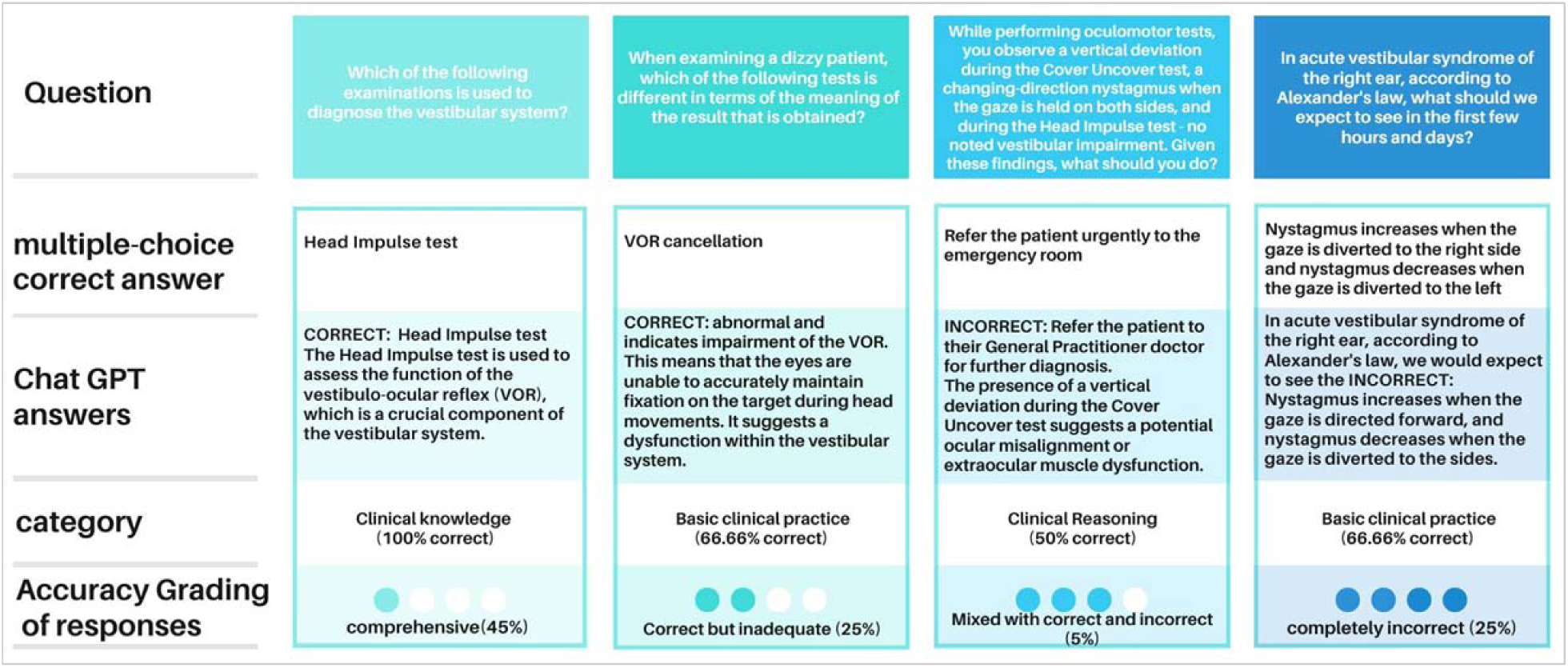
Examples of ChatGPT prompts for multiple-choice questions, and expert-graded ChatGPT responses.

### Intercorrelation among experts

A Spearman correlation test was conducted to assess the intercorrelation among the judges by examining the grading of each pair of experts. The correlation coefficients ranged from 0.68 to 0.76, indicating a moderate to strong positive association between the judgments of the experts (rs = 0.68 to 0.76, p < 0.05).

## Discussion

In this study, we present a novel analysis of ChatGPT’s and Google Gemini’s performance in the domain of vestibular rehabilitation. Our study shows clinicians are better at solving complex clinical questions that demand clinical reasoning abilities than ChatGPT and Google Gemini. In addition, phrasing the question correctly and carefully is crucial if a clinician chooses to consult with AI technology ChatGPT outperformed Google Gemini on VKT with significant and intriguing divergences in their responsiveness across categories. In the Clinical Knowledge category, both AI models excelled, however, they struggled in the Clinical Reasoning category. This difference in performance indicates that while chat-based platforms like ChatGPT and Google Gemini can be effectively employed for topics related to Clinical Knowledge, they may face limitations in dealing with more complex scenarios encountered in the Clinical Reasoning category. This highlights the fascinating differences between clinicians and artificial intelligences, and the comparative level of complex thinking needed in the field.

Our research findings reveal that ChatGPT and Google Gemini had a 70% and 60% accuracy rate, respectively, in answering multiple-choice questions in VR. This is in line with previous studies of ChatGPT’s performance in medical exams such as the USMLE (60%)^13^, AHA ACLS (76.3%),^12^ and Medical Physiology Examination of Phase I MBBS (>75% of marks)^20^. Similar to this, ChatGPT answered properly to 63% of the questions in the vestibular system category of another study that used quiz-style questions from the German Society of Oto-Rhino-Laryngology platform (n=95 accurate vs. n=57 false).^21^ Our study confirms previous research, highlighting the consistent performance of AI in specialized medical fields, as well as the important differences between two major AI chatbots.

In line with previous research, such as Smith et al. (2021)^22^ which assessed AI capabilities in neuro-ophthalmological diagnostics, our results also demonstrate that ChatGPT outperforms Google Gemini.^22^ This consistent finding has prompted us to delve deeper into the explanations ChatGPT provided for its responses. The frequency of errors in ChatGPT’s explanations in our study is particularly notable in the Clinical Reasoning category. Indeed, errors often occur when addressing questions that involve clinical reasoning and descriptions, which require a deep understanding of medical contexts rather than solely relying on clinical knowledge. For example, when providing a description of a patient who exhibits a down-beating nystagmus with a rightward rotation during Dix-Hallpike tests on both sides, the ChatGPT response concluded that the "most appropriate treatment choice would be: A positioning treatment for the right posterior canal by performing a Gufoni maneuver." While ChatGPT did accurately diagnose the right ear, it was wrong when it came to the identification of the canal and the appropriate treatment. ChatGPT’s performance typically decreased in questions requiring intricate descriptions of signs and symptoms.

An interesting finding was revealed from ChatGPT’s responses to post-treatment restrictions regarding prohibitions and restrictions. In this context, ChatGPT’s answers were found to be incorrect and based on outdated approaches and previously published articles^23^ available on the Internet. For example, when asked what recommendations should be given after a patient is successfully treated with BPPV concerning what should he or she should avoid doing at home, the Chat incorrectly responded: "Avoid lying on the side that was treated". ChatGPT’s responses were influenced by the abundance of information available online rather than by recent and reliable sources.^7^ ChatGPT, however, did provide accurate and current answers when the question was rephrased: In response to, “After you successfully treat a patient diagnosed with BPPV, what will be your post-treatment recommendations?”, ChatGPT’s response was, "Return to normal activity; you must not refrain from movement or restrict the sleeping position but should avoid climbing ladders or stools."

Thus, in making use of ChatGPT, careful consideration should be given to the impact of the wording of the question and presentation scenarios, as it could affect the quality of the ChatGPT response. Additionally, it is crucial to formulate questions carefully and be aware that ChatGPT’s responses may be influenced by the prevalence of information on the web rather than being based on updated, current and reliable sources. We ensured that no prompting or training was provided to the AI, by entering each question separately using the same chat.

It is evident that the utilization of AI for enhancing clinical decision-making will continue to expand. This growing trend highlights the necessity for effective collaboration between medical professionals and technology developers. With the rapid growth of medical knowledge, the integration of technologies like AI becomes crucial in enabling healthcare professionals to effectively apply this knowledge in their practice.

Health care education emerges as a captivating domain to explore, given the vast amount of information and diverse concepts that healthcare students are expected to comprehend.^24^ For example, in a recent editorial by Moons and Van Bulck (2023), they highlight the potential of ChatGPT in cardiovascular nursing practice and research. They further emphasize its ability to summarize large texts, facilitate the work of researchers, and assist in data collection, making ChatGPT a potentially valuable tool in health care practice.^25^

Our study, together with previous research, leads us to formulate the following recommendations regarding the specific cases where ChatGPT and Google Gemini can be utilized to expedite the learning process in clinical knowledge. Instead of investing time on reading and memorizing scientific literature updates and guidelines, ChatGPT and Google Gemini can provide a viable alternative. Indeed, a recent systematic review examined the potential applications of LLMs in healthcare education. The review highlighted several benefits of ChatGPT and Google Gemini in healthcare education, such as enhanced personalized learning experiences and an emphasis on critical thinking and problem-based learning.^26^ Likewise, instead of sifting through every necessary article or available data, ChatGPT and Google Gemini can diminish the workload by retrieving only the most relevant information, providing that the user ensures that the uploaded data is up-to-date and is precise when prompting the AI.

Our research confirms previous findings, showing that ChatGPT and Gemini can improve learning by making information accessible and aiding information retrieval which are part of the abilities required at the basic levels of Bloom’s Taxonomy learning pyramid.^27^ Additionally, these tools facilitate the acquisition of foundational knowledge, potentially enabling deeper engagement with content and more complex learning tasks.

To determine to what extent ChatGPT and Google Gemini could provide accurate responses and reliable information, further research is needed to validate the efficacy of ChatGPT and Google Gemini in providing accurate answers in the medical domain, specifically in the context of VR. Furthermore, studies should be conducted to determine if ChatGPT’s and Google Gemini’s performance can be enhanced through techniques such as question repetition and integration of reliable medical literature. As emphasized by ChatGPT itself, the evaluation of its output by professionals remains crucial in ensuring the accuracy and completeness of the information provided. In our opinion, clinicians who wish to “consult” with ChatGPT and Google Gemini should simplify their questions and avoid clinical reasoning questions.

## Study limitations

This study acknowledges the following limitations. First, there is a lack of a standardized questionnaire to assess knowledge and clinical reasoning in vestibular rehabilitation. To address this, we developed and validated a new questionnaire, the Vestibular Knowledge Test (VKT). While the VKT employs a multiple-choice format with 20 questions, its development process ensured content validity through expert review and pilot testing with vestibular rehabilitation professionals.

Second, the VKT focuses on three core vestibular rehabilitation categories: (1) Clinical Knowledge, (2) Basic Clinical Practice, and (3) Clinical Reasoning. This may not encompass all relevant knowledge areas. However, these categories represent foundational aspects of vestibular rehabilitation practice. Future iterations of the VKT could be expanded to include additional knowledge domains.

Third, the initial development and administration of the VKT occurred in Hebrew, with subsequent translation to English. While professional terminology in vestibular rehabilitation is predominantly English, the translation process underwent rigorous review by bilingual experts to minimize linguistic bias. This potential bias was further mitigated by acknowledging it as a limitation and by ensuring a clear and consistent translation.

## Conclusions

The ChatGPT and Google Gemini platforms are effective tools that can be used to obtain information and answer questions related to a variety of fields, including VR. However, it’s important to recognize that these platforms have limitations. They may provide inaccurate answers, particularly in complex areas like clinical VR reasoning. Careful phrasing and choosing the right words when prompting these AI tools are crucial for getting the most accurate and relevant results. Medical professionals, especially, should use these tools with caution and be aware of their limitations.

## Data Availability

All data produced in the present study are available upon reasonable request to the authors

## Acknowledgments

We would like to express our gratitude to Prof. Avi Shupak, Prof. Yahav Oron, Dr. Amit Wolfovitz, as well as the physical therapists and physical therapy students who participated in the study.

## Ethics approval and consent to participate

The study was approved by the Ethics Committee for Non-clinical Studies at Bar Ilan University in Israel. The ethics form was signed by the committee head and the date of approval was 21 May 2023.

